# Association between recent COVID-19 diagnosis on depression and anxiety symptoms among slum residents in Kampala, Uganda

**DOI:** 10.1101/2022.12.28.22284012

**Authors:** Solomon T. Wafula, Lesley L. Ninsiima, Hilbert Mendoza, John C. Ssempebwa, Florian Walter, David Musoke

## Abstract

**Background:** An increase in mental health problems has been reported since the beginning of the COVID-19 pandemic. However, little is known about the prevalence of depressive and anxiety disorders, and how recent COVID-19 diagnosis may influence risk of these conditions especially in low-income settings. In this study, we assessed the association between recent COVID-19 diagnosis and depressive and anxiety symptoms among residents in an urban slum setting in Uganda.

**Methods:** A cross-sectional study was conducted among 284 individuals in a slum settlement in Kampala, Uganda between April and May 2022. We assessed generalized anxiety and depression symptoms using two validated questionnaires. We collected data on sociodemographic characteristics, and self-reported recent COVID-19 diagnosis (in the previous 30 days). Using a modified Poisson regression, adjusted for age, sex, gender and household income, we separately provided prevalence ratios and 95% confidence intervals for the associations between recent COVID-19 diagnosis and depressive and anxiety symptoms.

**Results:** Overall, 33.8% and 13.4% of the participants met the depression and generalized anxiety screening criteria respectively. People with recent COVID-19 diagnosis were more likely to be depressed (53.1%) than those with no recent diagnosis (31.4%). Participants who were recently diagnosed with COVID-19 reported higher prevalence of anxiety (34.4%) compared to those with no recent diagnosis of COVID-19 (10.7%). After adjusting for confounding, recent diagnosis with COVID-19 was associated with depression (PR= 1.60, 95% CI 1.09 – 2.34) and anxiety (PR = 2.83, 95% CI 1.50 – 5.31).

**Conclusion:** This study suggests an increased risk of depressive symptoms and GAD in adults following a COVID-19 diagnosis. We recommend additional mental health support for recently diagnosed persons. The long-term of COVID-19 on mental health effects also need to be investigated.

## Introduction

In 2020, the World Health Organization (WHO) declared COVID-19 a pandemic and has since had detrimental effects on population health and well-being worldwide [1]. The uncertainties and unpredictable nature of the pandemic resulted in individuals experiencing various forms of mental health problems such as depression, anxiety, psychosocial dysfunction, dissociation disorders, substance abuse, and insomnia. It was projected that these effects would be most severe in low-resourced settings due to constraints related to weak health systems with limited access to mental health services [2].

Low-resourced settings such as slum residences comprise of informal dwelling units which are poorly constructed, crowded with poor sanitary conditions and environmental pollution [3, 4]. Slum dwellers usually have poor physical and economic access to health care including mental health care [5]. Most informal settlements comprise an assortment of individuals (e.g., low-income earners, asylum seekers, refugees), who are at high risk of mental health problems due to multiple stressors and this may be further aggravated by the COVID-19 pandemic [2]. Due to unfavourable conditions of unstable income sources, high dependence on daily casual jobs, and threatened livelihoods and food security [6, 7], slum dwellers may be at elevated risk of experiencing mental health challenges. A constellation of individual, social and cultural factors that contribute to psychological distress in informal settings have been highlighted and include trauma, violence, community insecurity, and healthcare barriers [8]. Mental health among informal settlement dwellers is understudied at large, including both before and during the COVID-19 pandemic[9]. Available evidence suggests that psychological distress during the pandemic has been high in various parts of Africa, For example, in a phone-based study in urban areas of Burkina Faso, Ethiopia, and Egypt, 28% of participants had symptoms of mild, moderate, or severe psychological distress [10].

Evidence from recent epidemics such as SARS, MERS and Ebola poignantly illustrate the psychosocial impacts of the epidemic and related restrictions. During these epidemics, the fear of infection or death, helplessness, depression, anxiety, social isolation, and stigma are the most reported psychological manifestations [11]. For example, approximately 15% of individuals quarantined due to SARS epidemic in the Canadian city of Toronto showed signs of depression and post-traumatic stress disorder [12]. Similarly, higher prevalences of psychological effects such as fear, anxiety and depression were reported during the 2013-2016 West Africa Ebola outbreak [13]. Whereas several studies on the effect of COVID-19 on mental health have been conducted elsewhere, there was limited evidence regarding the effect of recent diagnosis with COVID-19 on depression and anxiety, especially among socioeconomically deprived populations such as slum dwellers. To date, little is known about the burden of mental health conditions disease in urban slums and related research among slum residents in Uganda and globally [9]. We, therefore, set out to investigate the association between recent COVID-19 diagnosis and depressive and anxiety symptoms among residents in an urban slum in Uganda.

## Methods

### Study design and setting

This study used cross-sectional, population-based data from adults (at least 18 years of age) residing in Bwaise; an urban slum setting in Kampala, Uganda from April to May 2022. Bwaise is one of Kampala’s most densely populated slums, distinguished by mostly informal and substandard housing and small-scale businesses. It has a high population density, crowded households, and low socioeconomic status. The study population consisted of adults who had resided in the area for at least three months.

### Sampling procedures

A sample of 284 adults from households in the Bwaise slum participated in the study. Sample size was calculated for the main study[14]. A representative sample of households were selected from the three parishes in Bwaise. Within each parish, one zone was selected randomly by ballot method, and at least 93 households were selected from each zone using systematic sampling, with a sampling interval obtained by dividing the number of households (based on records from the local office (LC)) by the number of households needed in each zone (i.e., divided by 93). Sampling and interviews started at the LC office and then north direction then clockwise until the target sample size was realised for each zone. A selected household was replaced by the next household if the original household had no eligible respondents or did not consent.

### Study procedures and measures

We organized face-to-face questionnaire interviews with participants through a consenting process by experienced and trained research assistants. Research assistants with a background training in health sciences captured participant responses on KoboCollect; a mobile data collection app (https://www.kobotoolbox.org/). We collected information on the exposure (recent COVID-19 diagnosis (in the previous 30 days) and having experiences (diagnosis or death) of a family member or close friend in the previous 30 days), as well as on covariates; the socio-demographic characteristics (age, marital status, income, education level, duration of stay), substance use in the last 30 days (alcohol, smoking, marijuana).

The primary outcomes were current depression and generalized anxiety disorder (within the preceding 2 weeks). Depressive symptoms were assessed using a validated 9-item Patient Health Questionnaire (PHQ-9) [15] with each item scored as 0 (not at all), 1 (several days), 2 (more than half the days) and 3 (nearly every day). Thus, the total score of the PHQ-9 can range from 0 to 27. As previously recommended [16], we used the cutoff score of 10, corresponding with at least a moderate level of depression. Anxiety was assessed using a 7-item validated Generalized Anxiety Disorder (GAD-7) tool to screen the presence of generalized anxiety disorder and assess its severity [17]. The recommended screening cutoff was ≥10, corresponding with at least a moderate anxiety level. Data collection tools were developed following a critical review of existing literature [18-21].

### Statistical analysis

Statistical analyses were performed using Stata 14 (Stata Corp, Texas, USA). Descriptive statistics such as frequencies and percentages were used for categorical data. For continuous data, we used medians and interquartile range. Participant characteristics by depression and anxiety status were compared using the Wilcoxon rank sum test for non-normally distributed continuous variables and the Chi-squared test for dichotomous variables. We also performed two separate multivariable modified Poisson regressions adjusting for age, sex, gender and household income [22] and producing prevalence ratios (PRs) and corresponding 95% confidence interval (95% CI) for the association between recent COVID-19 infection and depression and anxiety. All statistical tests were two-tailed, and statistical significance was assumed when a p-value was ≤ 0.05.

## Results

### Background characteristics of participants

In total, 284 participants participated in the study including responding to the PHQ-9 and GAD-7 questionnaires. Characteristics of all participants are described for depressive and anxiety disorders determined by PHQ-9 and GAD-7 cutoffs ≥10. Most participants were females 85.2% and had a median age of 29 years (IQR = 24,37). The median depression score based on PHQ-9 was 8 (6,10) while for anxiety, the median score was 6 (4,8) based on the GAD-7. The prevalence of depression was 33.8% and anxiety was 13.4%, which differed significantly depending on income (p<0.001) and recent positive COVID-19 diagnosis status (p<0.001) (Table 1).

**Table 1.**
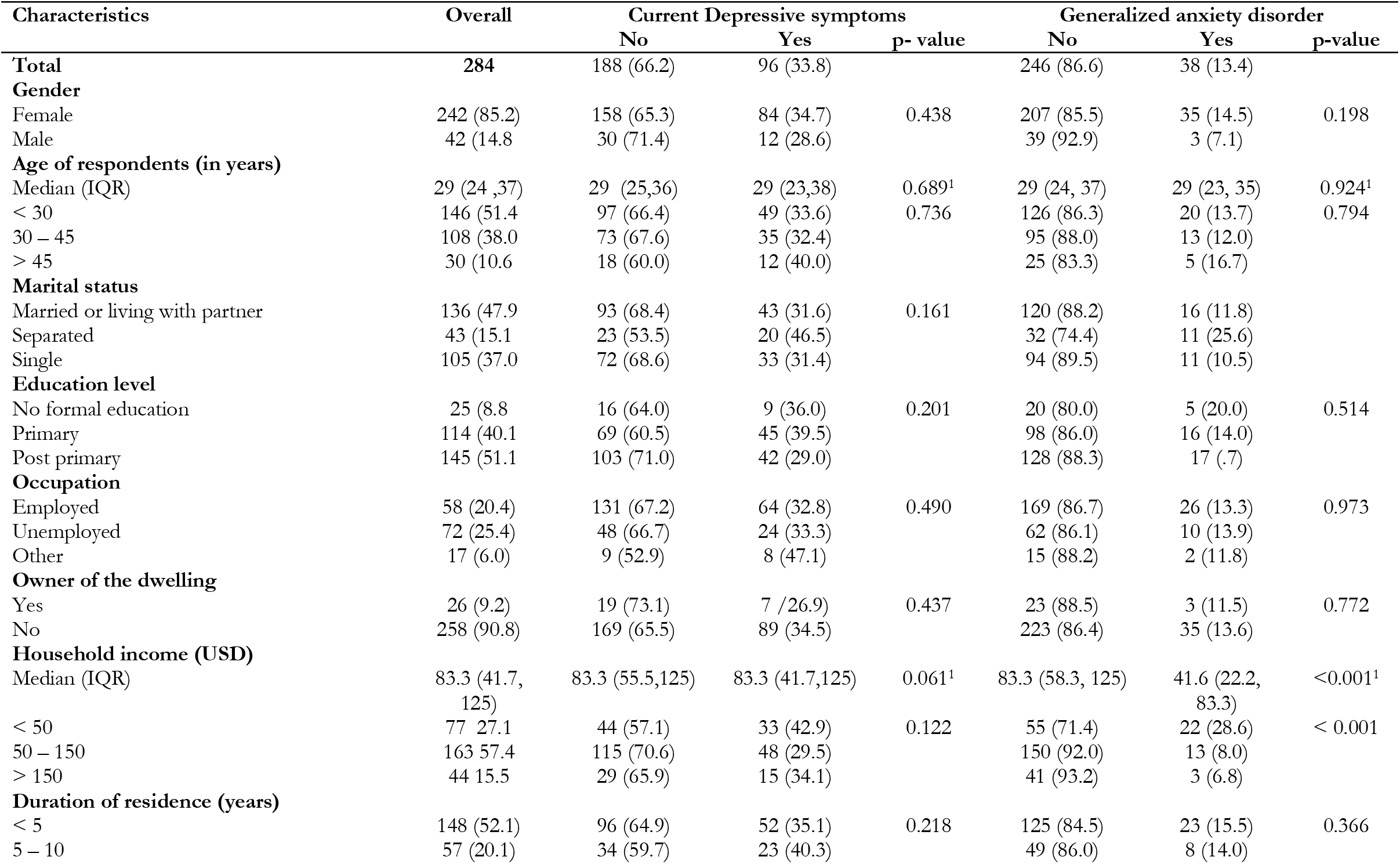
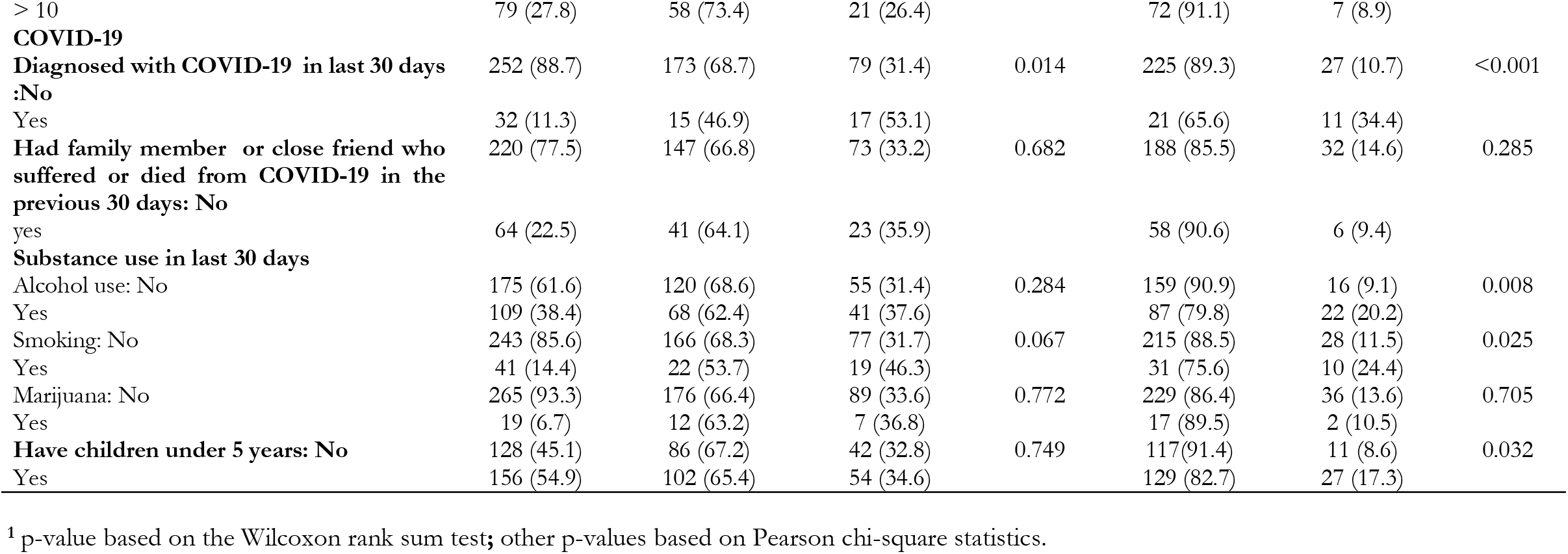
Characteristics of household participants.

### Association between recent COVID-19 diagnosis and depressive symptoms

In unadjusted analysis, there was a significant association between recent SARS-COV-2 infection with household income and smoking in households. After adjusting for age, gender and income, recent COVID-19 diagnosis was positively associated with current depressive symptoms (PR= 1.60, 95% CI 1.09 – 2.34) (Table 2).

**Table 2.** Association between recent COVID-19 diagnosis and depressive symptoms.

| Attributes | Current depressive symptoms |  |  |  |
| --- | --- | --- | --- | --- |
|  | Unadjusted analysis |  | Adjusted analysis |  |
|  | PR (95 % CI) | p-value | PR (95 % CI) | p-value |
| ***Sociodemographic characteristics*** |  |  |  |  |
| **Gender** |  |  |  |  |
| Female | 1 |  |  |  |
| Male | 0.82 (0.45 – 1.51) | 0.454 | 0.83 (0.49 – 1.39) | 0.470 |
| **Age of respondents (in years)** | 1.00 (0.98 – 1.02) | 0.872 | 1.00 (0.98 – 1.01) | 0.896 |
| **Marital status** |  |  |  |  |
| Married or with partner | 1 |  | 1 |  |
| Separated | 1.47 (0.86 – 2.50) + | 0.062 | 1.25 (0.81 – 1.92) | 0.312 |
| Single | 0.99 (0.63 – 1.56) | 0.975 | 0.92(0.62 – 1.36) | 0.665 |
| **Education level** |  |  |  |  |
| No formal education | 1 |  |  |  |
| Primary | 1.09 (0.54 – 2.24) | 0.752 |  |  |
| Post primary | 0.80 (0.39 – 1.65) | 0.464 |  |  |
| **Occupation** |  |  |  |  |
| Employed | 1 |  | 1 |  |
| Unemployed | 1.01 (0.69 – 1.49) | 0.937 | 0.98 (0.65 – 1.48) | 0.931 |
| Other | 1.43 (0.83 – 2.47) | 0.194 | 1.41 (0.78 – 2.55) | 0.253 |
| **Owner of the dwelling:** | 1.28 (0.59 – 2.76) | 0.459 |  |  |
| **Length of stay (years)** |  |  |  |  |
| < 5 | 1 |  |  |  |
| 5 - 10 | 1.14 (0.70 – 1.88) | 0.481 |  |  |
| >10 | 0.76 (0.46 – 1.26) | 0.201 |  |  |
| **Household income (USD)** |  |  |  |  |
| < 50 | 1 |  | 1 |  |
| 50 – 150 | 0.69 (0.48 – 0.98) | **0.036** | 0.80 (0.55 – 1.15) | 0.224 |
| > 150 | 0.80 (0.49 – 1.29) | 0.356 | 0.83 (0.51 – 1.36) | 0.457 |
| ***COVID-19 experiences*** |  |  |  |  |
| Diagnosed with COVID-19 in the last 30 days | 1.69 (1.01 – 2.86) * | **0.006** | 1.60 (1.09 – 2.34) | **0.016** |
| Family or close friend suffered or died from COVID-19 in the previous 30 days | 1.08 (0.67 – 1.73) | 0.679 |  |  |
| ***Psychoactive substance uses*** |  |  |  |  |
| Alcohol use | 1.20 (0.80 – 1.79) | 0.281 |  |  |
| Regular smoking | 1.46 (0.89 – 2.42) | **0.049** | 1.29 (0.85 – 1.96) | 0.225 |
| Marijuana or cannabis use | 1.09 (0.51 – 2.36) | 0.768 |  |  |
| **Have children under 5** | 1.05 (0.70 – 1.58) | 0.750 |  |  |

### Association between recent COVID-19 diagnosis and generalized anxiety disorder

In adjusted analysis, participants who had been diagnosed with COVID-19 within 30 days of the survey had a 2.8 times higher risk of anxiety compared to their counterparts (PR = 2.83, 95%CI 1.50 – 5.31). Higher monthly incomes (i.e. USD 50 – 150 (PR = 0.38, 95%CI (0.19 – 0.76) and USD> 150 (PR = 0.30, 95%CI (0.11 – 0.87)) were associated with lower risk of anxiety symptoms compared to those who earned less than USD 50. The likelihood of meeting anxiety screening criteria was 2.23 times higher among particular who reported regularly drinking alcohol compared to those with no/limited alcohol consumption (PR = 2.23, 95CI 1.12 – 4.42) (Table 3).

**Table 3.**
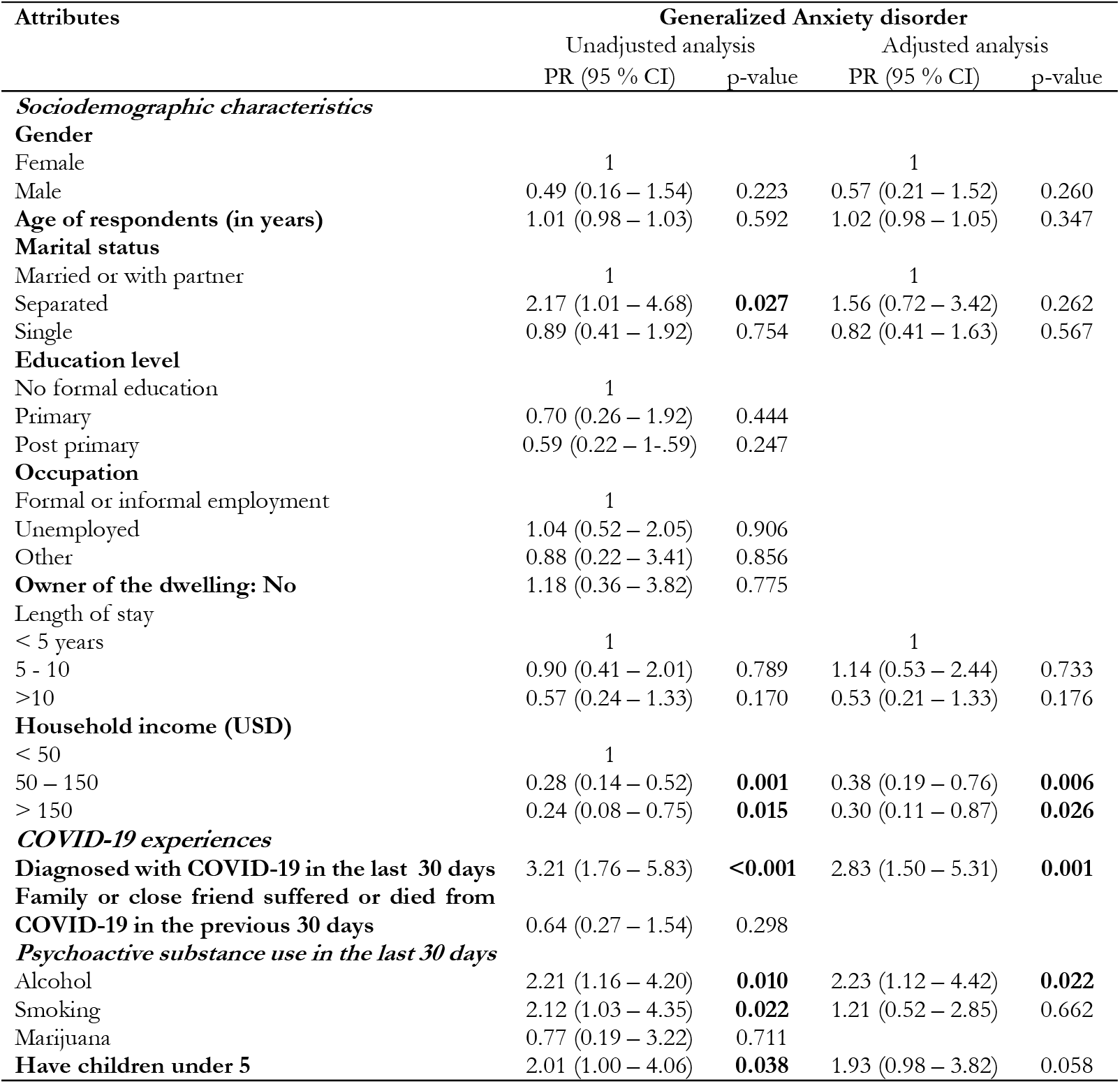
Association between recent COVID-19 diagnosis and anxiety disorder.

## Discussion

In this community-based study, we examined the prevalence of depressive symptoms and anxiety and their association with recent COVID-19 diagnosis among adult slum dwellers in Kampala, Uganda. Our findings highlight the challenges that individuals affected by COVID-19 face and are consistent with the larger literature on the impact of COVID-19 on mental health in the general population. In this study, a recent COVID-19 diagnosis was associated with both depression and anxiety. For instance, it increased the likelihood of depressive symptoms by 60%. For GAD, the prevalence was 2.8 times higher than for those who did not have a recent infection after adjusting for age, gender, household income and use of psychoactive substances.

Our study results revealed a high prevalence of anxiety and depression among participants (13.4% and 33.8%, respectively). High levels of depression and anxiety raise concerns about the capacity of mental healthcare systems to respond during the pandemic. The prevalence of depression in this study was higher than national pre-pandemic levels of 29.3%[23]. In addition to studies in the general adult population in Uganda, there is evidence that mental health was significantly affected by the COVID-19 pandemic even when not directly affected by the infection [24]. In the previous study, the prevalence rates were reported to be in excess of 80% for depression and 90% for anxiety. Although outcome measures are not directly comparable to our study, the reported rates appear worryingly high. As noted in that study, several limitations related to the data collection might have artificially increased the prevalence rates. However, the study underlines the significance of the mental health impact attributed to COVID-19 and related restrictions. Our findings support new evidence from other LMICs on the state of mental health during the pandemic. For instance, a recent study conducted in a socially and economically comparable setting in Kenya, reported elevated rates of depression (34.1%) and GAD (14.0%) [2] while in Bangladesh, the estimated prevalence of depression was 33.0%[25]. A further study from the United Kingdom[26] has compared pre- and post-COVID-19 rates of psychological distress and has shown that deterioration of mental health did not recover after COVID-19 restrictions were eased.

Further evidence indicates mental health problems among adults, especially during a health crisis, impact negatively on child developmental and behavioural outcomes[27]. The implication is that children by parents/guardians with such environments and backgrounds may be at elevated risk to experience various cognitive, behavioural, and emotional problems later in life if the problem is not addressed [28].

Although several studies have highlighted the elevated risk of mental health disorders and psychological distress during the COVID-19 pandemic, there seems to be limited research focusing specifically on how being recently diagnosed with COVID-19 affects individuals’ mental health. In our study, recently diagnosed (< 30 days) participants were 1.5 times and 2.8 times more likely to be diagnosed to have depression and anxiety respectively compared to their counterparts. In line with these findings, evidence from the United States (23) showed that experiencing a COVID-19 infection was associated with an increased risk of common mental disorders such as anxiety and depression in the acute phase of the infection. This is also in agreement with multiple other studies reporting an elevation in mental health problems following infection [29-32]. Our study is, to our knowledge, this is one of the first studies to investigate the influence of recent COVID-19 diagnosis on depression and anxiety in Uganda and reinforces the fact that the month following an infection seems to be associated more strongly with depressive symptoms and anxiety than experiencing a COVID-19 infection at any point in the past[33]. A possible explanation for these findings is that additional stressors related to the impact of COVID-19 infections such as financial problems, isolation from others, grief and worries about others could contribute to the experience of common mental health problems. However, studies have suggested that underlying biological mechanisms could also explain the increase in the prevalence of mental disorders [34, 35]. It is well established that the mechanisms, predominantly driven by the body’s immune response and associated cytokine activity can affect the neurological structures of patients. It is therefore reasonable to suggest that these changes could contribute to the observed effects on mental health. However, these mechanisms are not fully understood and require further research.

## Limitations

Although our study is the first to examine the association between recent COVID-19 diagnosis and depressive symptoms and GAD in an informal settlement in Uganda, it had several limitations. First of all, a recent COVID-19 diagnosis was self-reported (as told to participants by medical practitioners after tests) and we could not validate this. It is possible that participants misclassified themselves as not having a COVID-19 infection because they were not tested or diagnosed during the study period (especially if they were asymptomatic). Second, we only collected data on a limited set of confounding variables, which, coupled with the relatively small sample size, may have hampered the robustness of our findings because we were unable to account for all relevant confounders. Thirdly, we assessed mental health cross-sectionally which limits our ability to test changes in prevalence over time and whether the associations changed as time since diagnosis increased and also as the pandemic progressed. However, considering that our findings correspond with the wider literature we are confident that despite unmeasured confounding, this association would still hold even if we were to adjust for further covariates. Longitudinal studies may be needed in future to assess the effect of COVID-19 diagnosis on mental health over time.

## Conclusion

Overall, the study suggests an elevated risk for depressive symptoms and GAD among adults within 30 days following COVID-19 diagnosis. These findings suggest that additional support is required to address this burden and ensure that people receive mental /emotional treatment when necessary. Although we evaluated the effect of COVID-19 diagnosis on mental health within 30 days, long-term effects are possible; therefore, future research should evaluate the long-term consequences of COVID-19 infections in LMICS.

## Data Availability

All relevant data are within the manuscript and its Supporting Information files.

## Declarations

## Acknowledgements

We appreciate the local authorities for administrative support as well as the research assistants and the participants for without which this study would not have been possible.

## Contributors

STW, MH and DM conceived and designed the study, STW supervised data collection, LLN, STW, HM and FW analyzed the data and wrote the first draft of the manuscript. STW, HM, JCS, FW and DM provided a critical review of the manuscript. All authors contributed to the interpretation of the results, reviewed and editing of the manuscript and approved the final version for publication.

## Data availability statement

All relevant data are available from within the manuscript as well as a supplementally information file.

## Ethical consideration

Ethical approval was obtained from Makerere University School of Public Health Higher Degrees Research and Ethics Committee (HDREC: Ref No. SPH-2021-99) and Uganda National Council of Science and Technology (UNCST: Ref No. SS996ES). The Kampala Capital City Áuthority gave permission for the study’s implementation, provided advise and support with planning of study activities. We followed principles of the Helsinki declaration and written informed consent was obtained from each participant.

